# The impact of heat stress on the human plasma lipidome

**DOI:** 10.1101/2024.06.10.24308716

**Authors:** Igor L. Estevao, Josh B. Kazman, Lisa M. Bramer, Carrie Nicora, Ming Qiang Ren, Nyamkhishig Sambuughin, Nathalie Munoz, Young-Mo Kim, Kent Bloodsworth, Maile Richert, Justin Teeguarden, Kristin Burnum-Johnson, Patricia A. Deuster, Ernesto S. Nakayasu, Gina Many

## Abstract

The year of 2023 displayed the highest average global temperatures since it has been recorded—the duration and severity of extreme heat are projected to increase. Rising global temperatures represent a major public health threat, especially to occupations exposed to hot environments, such as construction and agricultural workers, and first responders. Despite efforts of the scientific community, there is still a need to characterize the pathophysiological processes leading to heat related illness and develop biomarkers that can predict its onset. Here, we performed a plasma lipidomic analysis on male and female subjects who underwent heat tolerance testing (HTT), consisting of a 2-h treadmill walk at 5 km/h with 2% inclination at a controlled temperature of 40°C. We identified 995 lipids from 27 classes, with nearly half of all detected lipids being responsive to HTT. Lipid classes related to substrate utilization were predominantly affected by HTT, with a downregulation of triacylglycerols and upregulation of free fatty acids and acyl-carnitines (CARs). We additionally examined correlations between changes in plasma lipids by using the physiological strain index (PSI). Here, even chain CAR 4:0, 14:0 and 16:1, suggested by-products of incomplete beta oxidation, and diacylglycerols displayed the highest correlation to PSI. PSI did not correlate with plasma lactate levels, suggesting that correlations between even chain CARs and PSI is related to metabolic efficiency versus physical exertion. Overall, our results show that HTT has a strong impact on the plasma lipidome and that metabolic inefficiencies may underlie heat intolerance.

## Background

As the magnitude and frequency of extreme global temperatures continue to rise, the public health burden of heat-related illness is projected to increase dramatically [1-3]. Workers from many occupations, including first responders, athletes, civil construction workers, and agricultural workers, are subjected to intense physical exertion in extreme temperatures (either heat or cold), chemical exposures, and psychological stressors. These exposures and stressors can cause major acute and chronic health problems, and in extreme cases, death [4, 5]. Our group has been performing multi-omics analyses to simultaneously study the mechanisms and biomarkers of these exposures and stressors [6, 7]. We investigated lipidomic responses to acute physical exertion to mimic field conditions in a cohort of wildland firefighters. Our results identified candidate biomarkers of strenuous exercise responses. We demonstrated that intense physical exertion alters omic profiles indicative of an increased susceptibility to respiratory infections and immune responses [6].

Heat exposure increases the incidence of heat-related illnesses, which are associated with exacerbated chronic illness, increased hospital admissions and risk of death [4, 5]. Excessive heat can denature proteins, destabilize lipoproteins, and liquefy cellular membranes, which can ultimately lead to tissue death and organ failure [8, 9]. The compensatory physiological responses to heat stress include increases in metabolism, cardiac output, blood flow to the skin, and sweat rate [8]. Once the body’s thermoregulatory mechanisms become overwhelmed, a spectrum of heat-related illnesses can emerge, ranging from heat cramps to heat exhaustion and life-threatening conditions, such as heat stroke [10, 11]. The response to heat appears heterogeneous across populations and influenced by factors, such as body mass index (BMI), race/ethnicity, aerobic conditioning, and genetics [12-15]. However, the mechanisms and factors contributing to heat intolerance are largely unknown and understudied. Early studies in the field of exercise physiology allude to a role of increased glycolysis and decreased lipid metabolism in heat intolerant individuals [16], where relative workload demands (oxygen consumption) are reduced in heat tolerant individuals during acute heat exposure [17]. Aside from these studies, animal studies point to genetic influences [18, 19]. Recent human transcriptomic studies revealed differentially expressed genes encoding proteins involved in unfolded protein responses, energy metabolism, and immunity [9, 20]. To mitigate the health impacts of extreme heat in the public, we urgently need to better understand the molecular mechanisms of heat intolerance and identify their predictive biomarkers.

As the physicochemical properties of lipids are highly affected by high temperatures [21] and previous studies allude to changes in lipid oxidation and abundance in heat intolerance [16, 22, 23], we studied the effects of heat on the composition of lipids in human plasma. We performed lipidomic analyses of plasma from individuals who underwent HTT and analyzed changes in lipid profiles following acute HTT. We also investigated the heterogeneity of this response by sex and correlations between individual lipid species with the individual’s exertional heat tolerance.

## Material and Methods

### Experimental design

Subjects were recruited under one of two studies to investigate physiological responses and clinical utility of a standardized HTT among recruits and service members referred for evaluation of heat tolerance. Subject eligibility criteria included: 18-45 years of age; non-obese (waist circumferences <39.4 in); non-hypertensive (resting blood pressure < 140/90 mmHg); non-pregnant/lactating; free of cardiovascular disease; free of glucose or blood-pressure medications; not being treated for mental health disorder; no personal or family history of malignant hyperthermia. For subjects with prior exertional heat illness, testing had to occur at least six weeks after their last episode.

The study consisted of a baseline evaluation and of a HTT on separate days. The baseline evaluation included a demographic/medical history questionnaire, anthropometric assessment, and VO_2_ Peak testing. Percentage body fat was assessed using the InBody 720, a multipoint bioelectrical impedance analyzer (InBody USA, Cerritos, CA). For the VO_2_Peak test, subjects underwent a 5 min warmup walk at 5.0 km/h and 2.0% incline, while wearing a Polar heart rate monitor (Polar USA Inc, Lake Success, NY) and a mask to measure expired respiratory gasses using open-circuit spirometry (Oxycon Mobile portable system, Viasys Healthcare Inc, Yorba Linda, CA). Next, the treadmill went to 0.0% incline, and they ran at a constant speed between 7.7 and 13.7 km/h (exact speed depending on warm-up heart rate). The incline then increased by 2.5% every 2 min until exhaustion or VO_2_ plateau [24].

For the HTT, participants arrived in the morning, and were dressed in shorts and a t-shirt. Women were tested between days three and nine of their follicular phase. Urine specific gravity had to be under 1.02 to ensure adequate hydration at baseline. Next, the HTT consisted of walking for 2 h at a pace of 5 km/h at 40 °C and 40% relative air humidity as described previously [25]. Real-time physiological measures included heart rate (HR) and core body temperature using a 10 cm rectal thermocouple (Type T Thermocouple with Thermes WiFi, Physitemp, Clifton, NJ). Participants were allowed to drink water *ad libitum*, up to one liter per hour.

The primary output from the HTT includes maximal HR and core body temperature during the last 5 minutes of walking. We also computed the PSI, based on change in HR and core temperature over the HTT [26]. PSI was used as the primary HTT outcome as it provides a continuous measure of both exercise and heat strain. The PSI equally weighs change in core temperature and HR over the test through an upper limit of 39.5 °C and 180 BPM, respectively. PSI ranges from 0 to 10, and values can be classified as minimal (0-2), low (3-4), moderate (5-7), high (7-8), and very high (9-10) [26]. The HTT protocol typically elicits a PSI level of around 4 or 5 [27], which is consistent with HTT being a moderately difficult heat challenge. Volunteers’ blood was collected pre- and post-exercise by phlebotomy and immediately placed on ice. The post-exercise sample was collected within 10 min after the test. Blood was drawn into EDTA-tube (Vacutainer, BD, Franklin Lakes, NJ, USA), mixed by inverting the tube 10 times, and immediately placed on ice. Plasma was then separated by centrifugation at 1000 ×g for 10 min at room temperature. Separated plasma was visually inspected for coagulation and hemolysis, collected into 0.5-mL aliquots, and stored within 1 h of blood draw at -80 °C for lipidomics analysis.

### Sample extraction

Plasma samples were extracted with the simultaneous metabolite, protein, and lipid extraction (MPLEx) protocol [28]. Samples (50 μL) were loaded into 2-mL Sorenson MµlTI™ SafeSeal™ tubes (Sorenson BioScience, Inc., Salt Lake City, UT, USA) with 10 μL SPLASH mix (in methanol) (Avanti Polar, Alabaster, AL, USA), and with 50 μL of a metabolite internal standard mix (1 mg/mL in water each ^2^H_4_-malonic acid, ^2^H_4_-succinic acid, ^2^H_5_-glycine, ^2^H_4_-citric acid, ^13^C_6_-fructose, ^2^H_5_-tryptophan, ^2^H_4_-lysine, ^2^H_7_-alanine, ^2^H_35_-stearic acid, ^2^H_5_-benzoic acid, ^2^H_15_-octanoic acid). Then, 390 μL of 2:1 (v:v) chloroform/methanol solution was added and samples were vortexed vigorously and incubated on an ice block for 5 min. Lipids, metabolites, and proteins were extracted into different phases by centrifugation at 12000 ×g for 10 min at 4 °C. The lipid layer (lower phase) was transferred into autosampler vials, dried in a vacuum centrifuge, and stored at -80 °C until liquid chromatography-tandem mass spectrometry (LC-MS/MS) analysis. The metabolite fraction was dried and incubated with 20 μL of methoxyamine in pyridine (30 mg/mL) for 90 min at 37 °C with shaking. The samples were derivatized with 80 μL of N-methyl-N-(trimethylsilyl)trifluoroacetamide with 1% trimethylchlorosilane for 1 h at 37 °C with shaking, and analyzed by gas chromatography-mass spectrometry (GC-MS).

### Gas chromatography-mass spectrometry

Derivatized metabolite fractions were analyzed on an Agilent GC 7890A using a HP-5MS column (30m × 0.25mm × 0.25μm; Agilent Technologies, Santa Clara, CA, USA) coupled to a single quadrupole MSD 5975C (Agilent Technologies, Santa Clara, CA, USA). One microliter of sample was injected in splitless mode with the GC oven preconditioned to 60 °C. This temperature was maintained for 1 min after injection, followed by an increase to 325 °C at a rate of 10 °C/min and a 10-min hold at 325 °C. A fatty acid methyl ester standard mix (C8-28) (Sigma-Aldrich, Saint Louis, MO, USA) was run in the instrument at the beginning of each batch for retention time calibration purposes. Retention time calibration and spectra deconvolution of GC-MS files were done using Metabolite Detector (version 2, Technical University, Braunschweig, Germany) [29]. Metabolites were identified by matching against the FiehnLib library [30] or the NIST17/Wiley 11 GC-MS spectral databases.

### Liquid chromatography-tandem mass spectrometry

Extracted and dried lipids were resuspended in 200 µL of 10% chloroform and 90% methanol. A total of 10 µL were loaded into a guard column (Acquity UPLC CSH C18 VanGuard Pre-column, 130 Å, 1.7 µM, 2.1 mm x 5 mm, Waters). The separation was carried out in a reverse phase column (Acquity UPLC CSH C18 column, 130 Å, 1.7 µM, 3.0 mm x 150 mm, Waters) connected to a Waters Acquity UPLC H class system (Waters Corp), maintained at 42 °C, over a 34 min gradient (mobile phase A: 40:60 (v:v) acetonitrile:water containing 10 mM ammonium acetate; mobile phase B: 10:90 (v:v) acetonitrile:isopropanol containing 10 mM ammonium acetate), at a flow rate of 250 µL/min with the following gradient profile (min, %B) 0, 40; 2, 50; 3, 60; 12, 70; 15, 75; 17, 78; 19, 85; 22, 92; 25, 99; 34, 99). Data were collected on an orbitrap mass spectrometer (Orbitrap Velos Elite, Thermo Fisher Scientific) in both positive and negative ionization modes. The MS1 scans were acquired at a range of m/z 200–2000. The top 4 most intense parent ions were selected for alternating high-energy collision and collision-induced dissociations, with an activation Q value of 0.18 and 35 normalized collision energy for collision-induced dissociation, and a normalized collision energy of 30 for high-energy collision dissociation.

### Lipid identification and quantitative information extraction

Raw LC-MS/MS files were converted to ABF format using Analysis Base File Converter (2011-2023 Reyfycs Inc.) and uploaded into MS-DIAL ver. 4.9.221218. MS-DIAL parameters were set for LC-MS/MS soft ionization and centroid data for MS1 and MS2. MS1 tolerance was set at 0.02 Da and MS2 at 0.025 Da. The minimum peak height was set at 10000. MS2 was deconvoluted and had chromatogram noise reduced with an abundance cutoff set as 5 in amplitude. The included adducts were [M-H]^-^, [M-H_2_O-H]^-^, and [M+Cl]^-^ for negative mode, and [M+H]^+^, [M+NH_4_]^+^, [M+Na]^+^, and [M+K]^+^ for positive mode. The alignment retention time was set at 0.25 min. All other parameters were kept as default values. Data curation for identification and individual annotation of molecules were performed manually based on the retention time relative to the SPLASH mix standards, carbon chain length, and MS2 spectra matched with the reference. One exception to this rule was the free fatty acids, which usually result in MS2 spectra without enough fragments for a confident identification. Because of its biological importance, we added the free fatty acid species for the statistical analysis and cross-validated it with the GC-MS data.

### Statistical analysis

Peak intensity values from MS-DIAL processing were exported for statistical data analysis. All data processing, quality control, and normalization were done using pmartR [31, 32]. Data was log2 transformed, and data quality was assessed by robust Mahalanobis distance to check for any outliers. Data was then normalized by global median centering. For each lipid, a mixed effects linear model was fit to account for the paired nature of the data; BMI, sex, timepoint, and interaction between sex and timepoint were included as fixed effects, and the subject was included as a random effect. The significance of lipid species abundances at pre- vs post-HTT was assessed using post-hoc comparisons, and a Benjamini-Hochberg p-value adjustment [33] was implemented. If the interaction term was significant post-hoc, comparisons of pre- vs post-heat tolerance for each sex were conducted with a Benjamini-Hochberg p-value adjustment applied. For each lipid, the correlation between heat tolerance index and lipid intensity change was assessed by the Pearson partial correlation test, while accounting for BMI [34]. Enrichment of regulated lipid classes were assessed by Fisher’s exact test. Lipid networks were built with Cytoscape v3.9.1 [35].

## Results

### Physiological responses to HTT

Overall acute changes in plasma lipids to HTT were measured in a cohort of 33 subjects, comprised of 26 male and 7 female subjects. The age of subjects spanned from 18 to 42 years. Subject anthropometric data are presented in Table 1. Changes in core temperature and HR were monitored throughout HTT as an indicator of heat tolerance. On average, baseline resting HR was 76 bpm, escalating to a final average HR of 123 bpm (Table 2). Baseline core temperatures commenced at an average of 36.9°C, rising to an average 38.0 °C at the end of HTT (Table 2). PSI is scored on a 1-10 scale where a higher numerical value indicates reduced heat tolerance. PSI scores ranged from a mild physiological strain of 2.18, and reached up to a high strain of 7.42, with overall values averaging 4.46, indicative of moderate strain. Notably, the variance (standard deviation) in core temperature and PSI were similar between female and male subjects, suggestive of similar levels responses to HTT between sexes. The average baseline and final HR for female subjects were 82 bpm and 141 bpm, respectively, while for male subjects, these values were 75 and 119 bpm (Table 2).

**Table 1.**
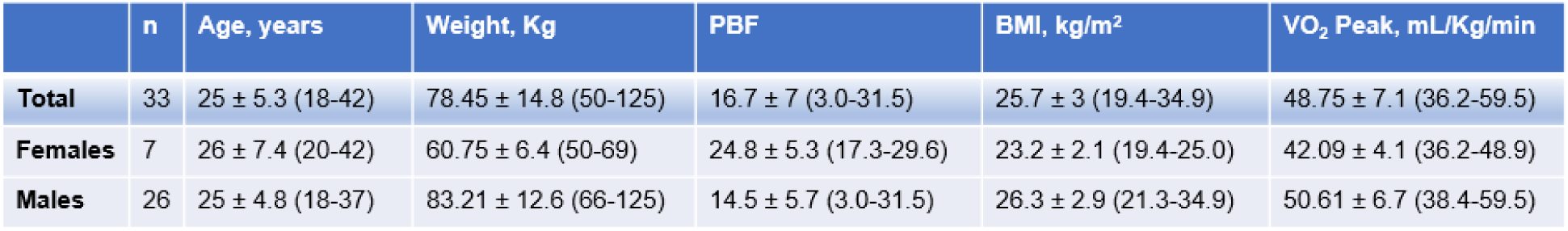
Baseline physical characteristics of subjects prior to exertional heat tolerance testing. VO_2_ Peak measurements were taken on 32 subjects (7 females and 25 males), whereas age, weight, percent body fat (PBF), and body mass index (BMI) were taken for 33 subjects. Data are represented as average, ± standard deviation and (range).

**Table 2.** Physiological responses to exertional heat tolerance testing. Physiological strain index (PSI), baseline core temperature, and final core temperature measurements were taken for 32 subjects (7 females and 25 males), whereas baseline heart rate and final heart rate were taken for 33 subjects. Heart rate is represented in beats per minute (bpm). Data are represented as average, ± standard deviation and (range).

|  | n | PSI | Baseline Heart Rate, bpm | Final Heart Rate, bpm | Baseline Core Temp (°C) | Final Core Temp (°C) |
| --- | --- | --- | --- | --- | --- | --- |
| **Total** | 33 | 4.5 ± 1.1 (2.2-7.4) | 76 ± 13.4 (39-107) | 123.4 ± 18.1 (88.4-158.5) | 36.9 ± 0.4 (36.3-37.9) | 38 ± 0.4 (37.1-38.9) |
| **Females** | 7 | 5.5 ± 1.1 (4.4-7.4) | 82 ± 21.8 (39-107) | 140.5 ± 24.1 (88.4-158.5) | 36.9 ± 0.4 (36.3-37.4) | 38.2 ± 0.4 (37.8-38.9) |
| **Males** | 26 | 4.2 ± 0.9 (2.2-5.7) | 75 ± 10.2 (54-98) | 118.8 ± 13.2 (97.4-143.8) | 36.9 ± 0.4 (36.4-37.9) | 37.9 ± 0.4 (37.1-38.7) |

### Changes in plasma lipids following HTT

Lipidomic analyses identified 995 lipids from 27 classes (Fig. 1A-B). Given a wide distribution of BMI in the data set (19.4-34.9 kg/m^2^), BMI was accounted for in statistical modeling. Further, given known sexual dimorphism in multiple metabolic processes [36], sex was included as a covariate in linear regression modeling. If a significant interaction between sex and timepoint was observed, changes in plasma lipids following HTT were assessed according to sex. After statistical analysis, 631 species had significant alterations, being 334 up- and 297 downregulated species, and 364 species were not significant across the cohort (Fig. 1A). We then performed an enrichment analysis to determine changes in plasma lipids based on class. Among lipids classes that predominantly function as phospholipids, phosphatidylcholines (PCs) were generally upregulated, while phosphatidylinositols (PIs), phosphatidylethanolamines (PEs) and lysophosphatidylethanolamines (LPEs) were downregulated following HTT (Fig. 1B). Of the sphingolipid lipid class, sphingomyelins (SMs) and dihexosylceramides (Hex2Cers) were upregulated (Fig. 1B). Among the sterols, cholesteryl esters (CEs) were enriched in upregulated lipid species with HTT (Fig. 1B).

**Figure 1.**
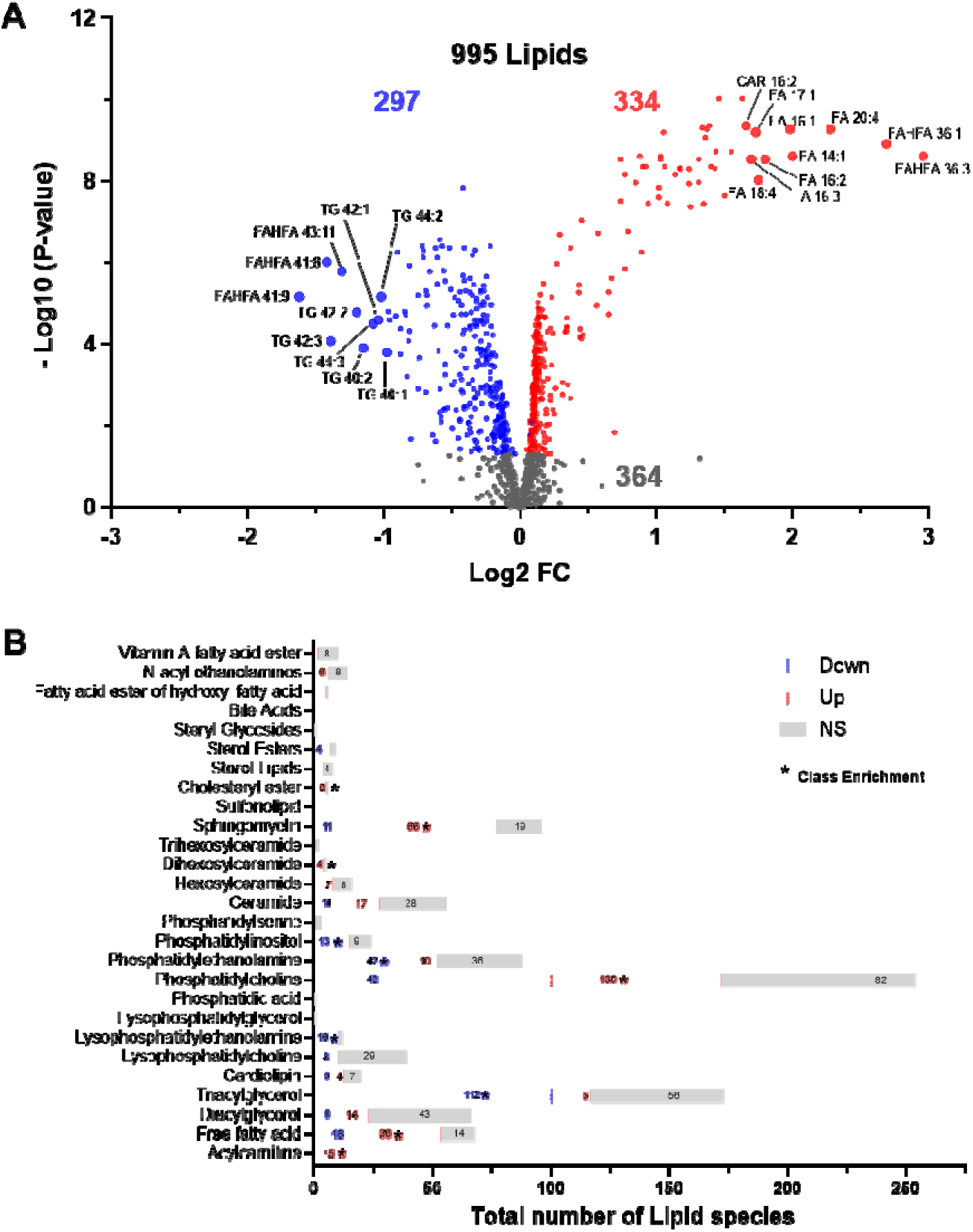
Effects of exertional heat stress on the plasma lipidome. **(A)** volcano plot displaying the number of differentially regulated plasma lipid species following heat tolerance testing, where each dot represents an individual lipid species. Upregulated species are indicated by red, downregulated in blue and non-significant (NS). **(B)** Distribution of up- (red) and down-regulated (blue) lipids species according to class. Class enrichment was assessed by Fisher’s exact test; p-values ≤ 0.05 are represented by asterisks (*).

Regarding lipid classes involved in energy metabolism, triacylglycerols (TGs) were largely downregulated with HTT (Fig. 1B). Among TG species, the majority that decreased were medium to short chain TGs (<45 carbon in length) that were largely saturated (indicated by a decreased number of double bonds) (Fig. 2A-B). This suggested that these TG species are preferentially consumed to sustain energetic demands during exertional heat stress. Diacylglycerols (DGs), which are intermediate degradation products of TG metabolism and involved in cellular signaling, displayed 14 species that increased and 9 that decreased with HTT (Fig. 1B). Both TGs and DGs are degraded into free fatty acids (FAs), which were enriched following HTT to include 36 upregulated species (Fig. 1B). As FAs often result in low confidence identifications in LC-MS/MS analysis, we performed a GC-MS to validate changes in their abundances. We found 5 FAs in common between the LC-MS/MS and GC-MS analysis: 14:0, 16:0, 16:1, 18:0, and 18:2. They all had similar patterns of upregulation after the HTT (Fig. S1), which validates the measurements of the LC-MS/MS analysis. In order for FAs to enter mitochondria for beta oxidation, they require conjugation to carnitines to form acyl-carnitines. CAR species were highly enriched, with all 15 detected species displaying an increase in abundance following HTT (Fig. 1B). The increase in CAR exemplifies the systemic metabolic demands of 2 hours of exertional heat stress.

**Figure 2.**
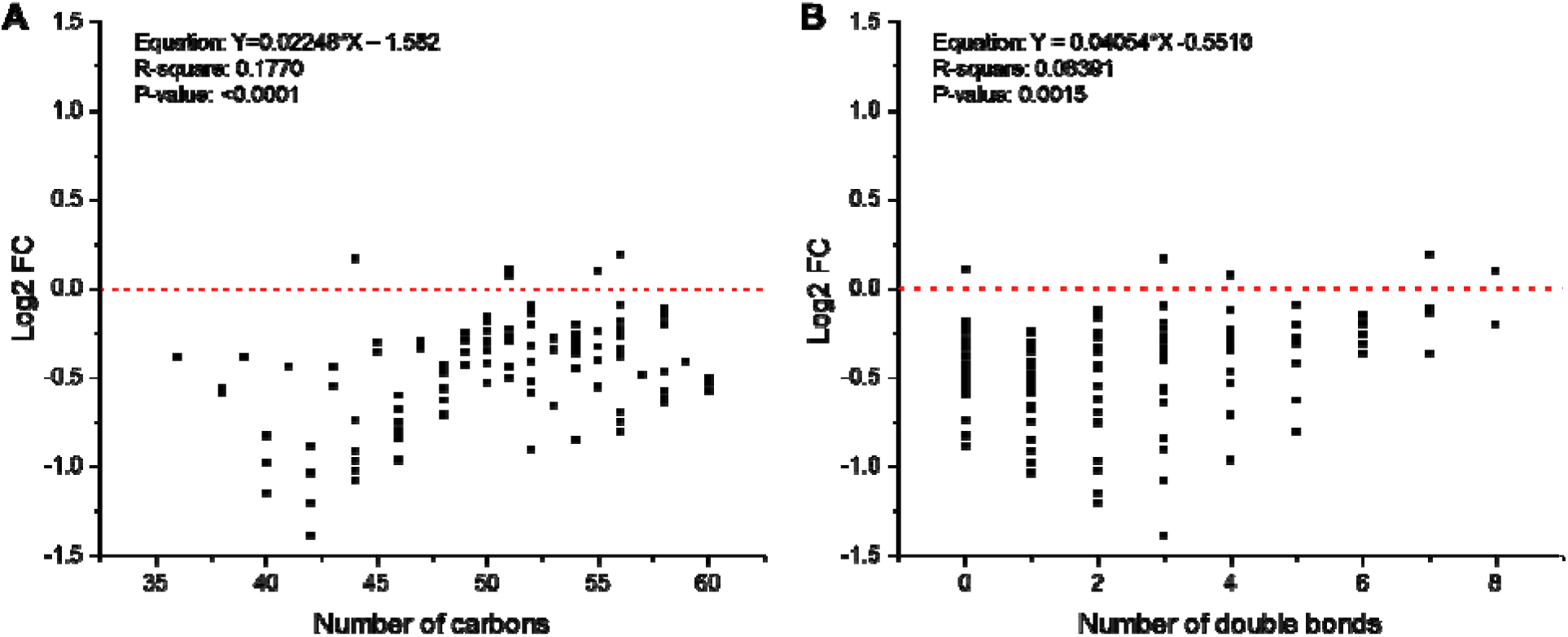
Changes in triacylglycerol species distribution following exertional heat tolerance testing. Changes in the triacylglycerol species distribution. A Linear regression curve was analyzed on log2 fold change values with exertion heat stress testing according to **(A)** carbon chain length and **(B)** double bond content.

We also investigated individual lipid species displaying the highest fold changes following HTT. Fifteen of the 20 most downregulated lipids were TGs, with the majority (70%) of the top 10 downregulated being short-chain saturated TG species, such as TG 40:1, 40:2 and 42:1 (Fig. 1A). This observation further supports that short-chain saturated TGs are preferentially utilized during HTT. We also observed that the relatively unexplored anti-inflammatory lipids, fatty acid ester of hydroxyl fatty acid (FAHFAs) [37] with long carbon chains and largely unsaturated (FAHFA 41:8, 41:9 and 43:11) were among the most downregulated lipid species after HTT, suggesting cellular uptake or signaling. Conversely, the shorter largely saturated FAHFAs 36:1 and 36:3 were among the most upregulated lipid species (Fig. 1A).

### Sex-dependent differences in lipid profile changes

We next investigated sex differences in the lipidomic response to HTT. A total of 117 lipid species exhibited a significant interaction with sex (Fig. 3A), indicating that the effects of HTT exposure differed in magnitude or direction between males and females. A total of 62 lipids were not regulated by the HTT, but had a significant sex interaction, reflecting basal sex differences in regulation of these lipids (Fig. 3A-B). Of the lipids with a significant by sex interaction, males and females shared 32 lipids that decreased and 10 that increased, suggesting that the magnitude, but not direction, of change differed by sex (Fig. 3A). For instance, PE 18:0_18:2 displayed similar reductions post HTT between sexes, but females had higher basal levels (Fig. 3C). Conversely, females had lower levels of CAR 16:0, but its abundance was raised to a similar final level in both sexes (Fig. 3D). Females displayed 10 lipids that increased and 3 that decreased following exertional heat stress that did not significantly change in males (Fig. 3A). This included PC 14:1_20:3, which increased only in females following HTT (Fig. 3E), whereas vitamin A fatty acid ester 21:0 decreased only in females (Fig. 3F). We next examined differences in lipid classes displaying a sex interaction.

**Figure 3.**
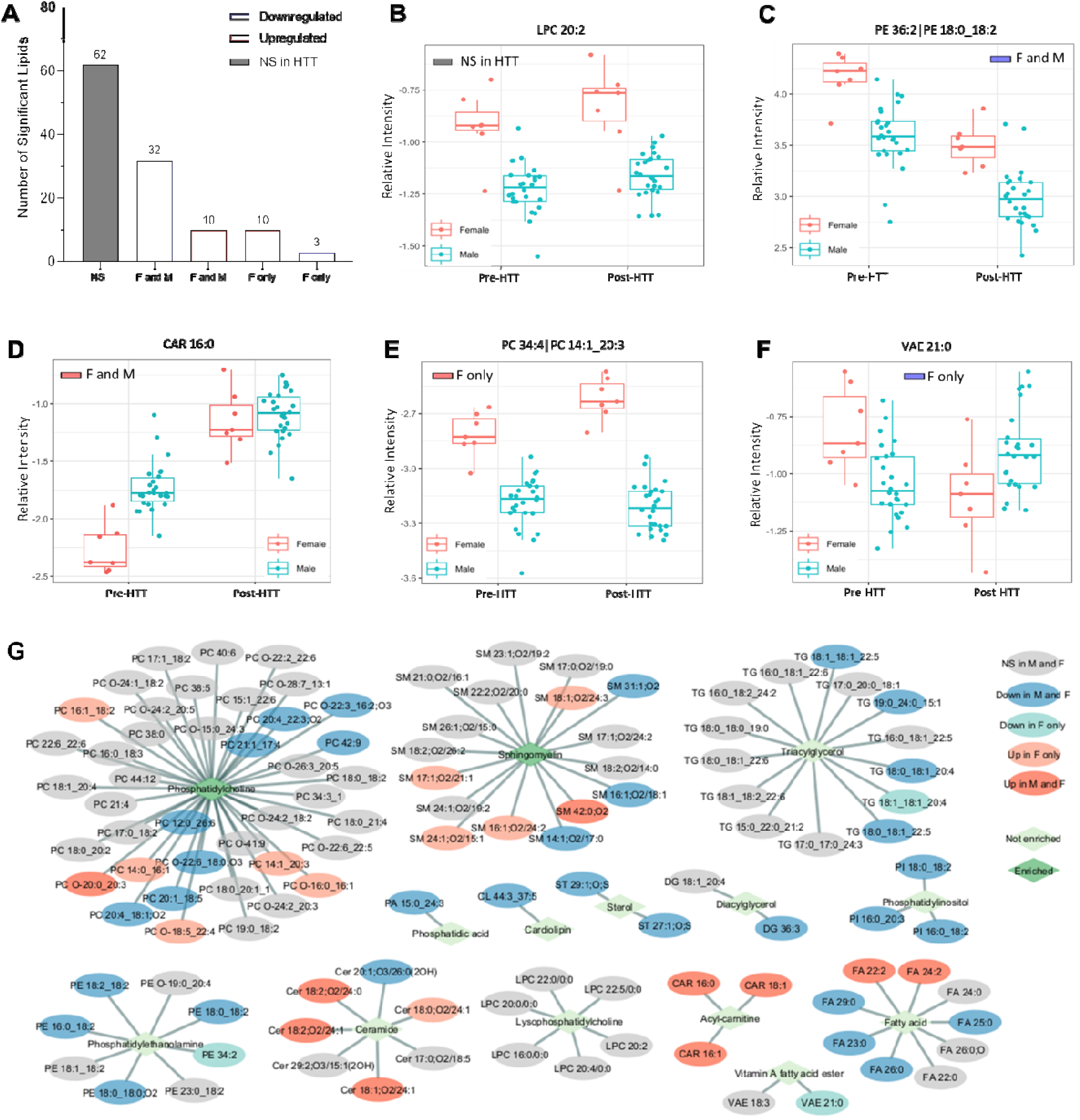
Differences in lipidomic responses to exertion heat stress testing between males and females. **A.** The number of plasma lipids displaying a by sex and timepoint interaction. Non-significant (NS) lipids (grey) represent those that are impacted by sex but did not change significantly with heat tolerance testing. Statistically significant down- and upregulated lipids that displayed a by sex and timepoint interaction are shown in blue and red, respectively. **B-F.** Box plots representing select lipid species from Fig. 3A. **G.** Network of individual lipid species and their corresponding lipid class that are significantly impacted by sex. Blue oval represents lipid species (LS) downregulated in both males and females, light turquoise color represents LS downregulated in females only, grey oval represents NS lipid species, light red represents upregulated LS in females only, and dark red oval indicates upregulated LS in males (M) and females (F). Light green rhombus is representative of a lipid class not significantly enriches, while a dark green rhombus is representative of enriched lipid classes.

Broadly, PC and SM lipids were the two primary lipid classes impacted by sex (Fig. 3G). PEs can be converted to PCs by methylation of their head group [38], whereas LPCs are formed by releasing FAs from PCs [39, 40]. Ceramides are converted to SMs by receiving the head groups from PCs [41-43], a class that displayed several of species impacted by sex (Fig. 3G), yet this lipid class did not display enrichment. Lipids related to energy metabolism, such as TGs, CARs, and FAs, had also numerous species impacted by sex and timepoint (Fig. 3G).

### Lipid correlates of exertional heat tolerance

We next sought to identify lipid species whose abundance profiles were associated with exertional heat tolerance. A higher PSI indicates reduced tolerance to exertional heat stress as indicated by a reduced ability to maintain HR and core body temperature during HTT [26]. We identified 26 and 35 lipids positively and negatively correlated with PSI, respectively. The lipid species were broadly distributed by class. Across sexes CAR species positively correlated with PSI (Fig. 4). The short chain acyl-carnitine, CAR 4:0, displayed the highest correlation to PSI, while long chain CARs, CAR 16:1 and 14:0 also displayed a significant correlation to PSI (Fig. 4). This paired with findings that short chain TGs, TG 8:0_9:0_11:0 and TG 8:0_10:0_15:2, were also positively associated with PSI. Conversely, medium to long chain TGs, such as TG 16:0_16:0_19:0;O2 and TG 18:0_18:1_20:4, were inversely correlated with PSI (Fig. 4). The TG degradation intermediates, DGs were also enriched with 8 species positively correlated with the PSI (Fig. 4). To determine whether the increase of intermediates of the energy metabolism was associated with higher physical exertion, we measured the lactate levels by GC-MS. No correlation was observed between PSI and change in plasma lactate levels or between PSI and final lactate levels (Fig S2A-B). In fact, only minor, not significant, changes in lactate levels were observed, an indication that the HTT is at relative low intensity for participants (Fig S2A-B). These results indicate that heat intolerance correlates with intermediates of lipid metabolism, but not necessarily with relative physical exertion during HTT.

**Figure 4.**
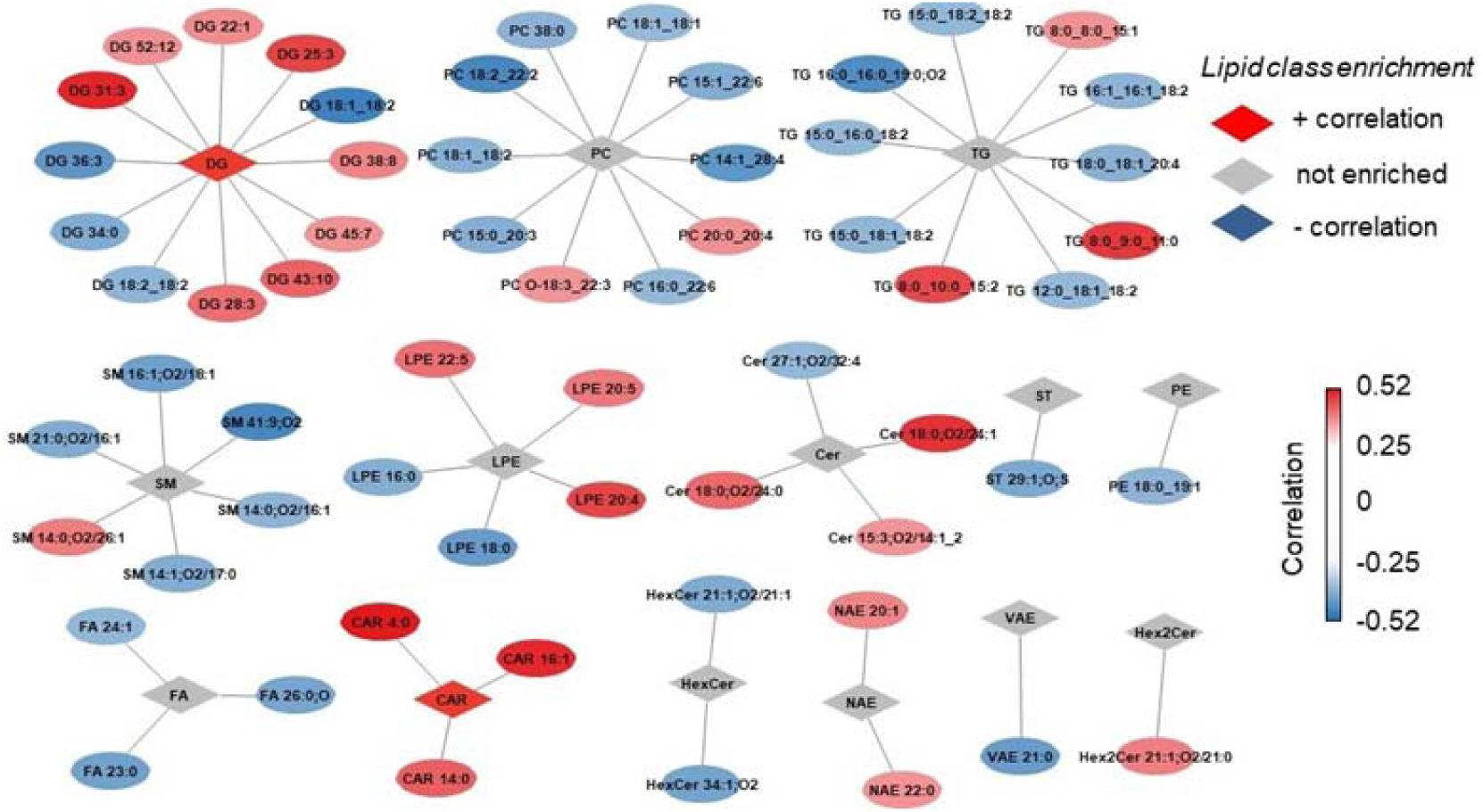
Computational network analysis of plasma lipid correlates to physiological strain index during heat tolerance testing. Visualization of positive (red rhombus) and negative (blue rhombus) enriched lipid classes. Correlation score in red/blue color saturation on lipid species represents fold change.

Additionally, sex-specific differences in lipids correlated to PSI were observed. In females, the short chain FA, 16:3 displayed the strongest correlation with PSI. Additional FA and CAR species such as FA 18:4 and CARs 18:1, 18:2 and 16:2 showed positive associations with PSI in females, but not males. Further, females displayed a strong negative association of the docosahexaenoic acid (DHA)-containing PE O-40:6 species (r= -0.95) [44] that was not observed in males. In males but not females, the anti-inflammatory FAFHAs 36:1,O and 36:3,O were negatively correlated to PSI. Additionally, in males 10 DG species were positively correlated to PSI, whereas only DG 36:1 was positively correlated to PSI in females. In males, but not females, DG 36:3 was negatively correlated to PSI. Interestingly DG 36:3 is reduced following sleep restriction [45], suggestive of its potential role as a biomarker of physiological stress.

## Discussion

Given rising global temperatures, exertional heat stress poses a significant public health burden that is magnified in occupational workers, such as athletes and military service members, with prolonged outdoor environmental exposures. Despite observed differential gene expression responses to environmental heat [7, 9, 18-20, 46, 47], there is limited data explaining the underlying mechanisms of exertional heat intolerance, let alone targetable biomarkers linked to increased risk of heat-related illness. Here, we complete a thorough analysis of exertional heat stress-responsive plasma lipids in male and female subjects. We show that two hours of exertional heat stress dramatically impacts the plasma lipidome. Notable responses include CAR, FA, SM and PC mobilization and reduced levels of short, largely saturated TG lipid species. We also demonstrate that the plasma lipidomic response to HTT differs between males and females and specific lipid classes correlate with reduced heat tolerance, to converge on an association between plasma levels of even chain CAR species 4:0, 14:0 and 16:1 and reduced heat tolerance. These findings were observed despite similar levels of physical exertion during HTT as indicated by plasma lactate levels. Together, our findings suggest that alterations lipid metabolism, among other changes in plasma lipid dynamics, underlie reduced tolerance to exertional heat stress.

Physiologically, a rise in core temperature or muscle temperature during exercise shifts substrate metabolism towards increased glycogen utilization and glycolysis [48, 49]. However, during heat acclimation substrate metabolism shifts to an increased reliance on lipids, likely in part facilitated by increased skeletal muscle FA uptake and utilization [16, 49, 50]. Here, we observed that HTT mobilized all detectable CAR species and broadly reduced short chain saturated TG species. Alterations in lipid classes such as TG, CAR, and DG highlight the intricate interplay between lipid metabolic systems in orchestrating the physiological response to thermal stress. We observed the TG intermediates, CAR, DG and ceramides, were correlated to heat intolerance in both sexes. CAR 4:0 displayed the highest correlation to PSI followed by DG 31:3, CAR 16:1 and ceramide 18:0;O2/24:1 This suggests an association between these lipids and heat intolerance. Incomplete beta oxidation results in accumulation of even chain CAR species and bi-products of amino acid catabolism to include CARs 3:0-5:0 [51].

Combined with an observed decrease in plasma TGs, this supports previous studies suggesting that glycogen sparing in the presence of increased lipid utilization fuels the increased metabolic demands of exertional heat stress in heat tolerant individuals [16, 22, 52]. It has been proposed that in insulin-resistance, which is characterized by mitochondrial dysfunction, fatty acyl-CoAs are diverted from transport to the inner mitochondria membrane and converted to DG and ceramide species [53]. Therefore, our findings suggest that incomplete beta oxidation, as indicated by an increase in plasma even chain CAR and DG species, may underlie heat intolerance.

The even chain CAR 4:0, 14:0 and 16:0 are associated with diabetes have been shown to impair insulin signaling and promote oxidative stress in cultured muscle cells [54]. Elevated levels of CAR 4:0 is observed in Parkinson’s disease models and genetic diseases of inborn errors of metabolism [55, 56]. Here, the production of protein carbonyls in response to these CARs is blocked by the antioxidant N-acetylcysteine, which suggest that elevated levels of these CAR species may promote oxidative stress. Future studies examining the role of these CAR species in pathophysiological responses to exertional heat stress are thus warranted.

We also observed sexual dimorphism in plasma lipid correlates of heat intolerance. Females displayed additional even chain CAR (CAR 16:2, 18:1, 18:2), TG (FA 16:3, 18:4) and DG species (DG 14:0_18:1) species that correlated with heat intolerance. Further, females displayed negative correlations of the DHA-containing PE O-40:6 species with PSI, suggesting a potentially protective role of DHA-containing lipids in exertional heat stress. In males, FAHFAs 36:1,O and 36:3,O which are suggested to have anti-inflammatory properties were negatively correlated to heat intolerance [57]. FAHFAs, branched fatty acid esters of hydroxy fatty acids, have been shown to have antidiabetic properties and attenuate LPS-mediated cytokine production. However, the molecular mechanisms by which they exert these properties are largely unknown [58]. It has been reported that physical activity promotes elevation of branched palmitic acid esters of hydroxy stearic acids (PAHSAs) in serum and adipose tissue. PAHSAs are FAHFA lipid mediators known for their insulin-sensitizing effects [59, 60]. It is therefore possible that FAHFA mobilize, or baseline levels are related to attenuated physiological response to exertional heat stress.

The current study is not without limitations. We included a cohort of relatively homogenous, physically fit adults. Given that heat intolerance is impacted by BMI, the responses in an obese cohort of subjects may differ [61]. Further, no subjects displayed a highly elevated PSI in response to HTT. An expanded cohort of subjects, including those of lower fitness, elevated BMI and greater number of female participants will help to better establish plasma lipidomic biomarkers. Additionally, HTT was used to elicit a thermoregulatory response to exercise, alongside a “plateau” in HR and core temperature among individuals who are heat tolerant [62]. While the American College of Sports Medicine recommends HTT for return to sport, some limitations of the clinical utility of HTT have been proposed as it involves exercising at a relatively low intensity [63]. Additionally, examination of lipidomic changes, in addition to standard clinical analytes of plasma lipids, like total TG content and non-esterified fatty acids, may help facilitate the clinical utility of this work. As such, examination of biomarkers in field-based settings may offer additional insight into the pathophysiological mechanisms of exertional heat intolerance.

To examine the role of varying physical exertion during HTT, we assessed changes in plasma lactate and found no correlations with PSI. This supports the notion that metabolic efficiency versus the relative level of physical exertion drives physiological responses to exertional heat stress. Future studies examining or controlling for other variables of physical fitness in addition to studying molecular changes at the myocellular level will help to elucidate potential mitochondrial or metabolic alterations associated with heat intolerance that are independent of physical fitness. In summary, our findings provide insight into the plasma lipidomic responses to exertional heat stress and elucidate candidate pathways regulating heat intolerance. Further investigation into the molecular mechanisms underlying these lipidomic changes promises to deepen our understanding of metabolic adaptation to environmental challenges and may hold relevance for the development of therapeutic interventions.

## Data Availability

Data will be uploaded and shared as a supplemental file to the text in the publication.

## Ethics approval and consent to participate

This study was performed in accordance with the World Medical Association, declaration of Helsinki and received approval from the Institutional Review Board (IRB) of the Uniformed Services University via protocol numbers: G191FY; FAM-81-3173-01.

## Authors’ contributions

ILE: Writing – original draft, methodology, data curation, investigation, data interpretation, review & editing. JBK: Investigation, methodology, writing, review & editing. LMB: Data analysis, writing, review & editing. CN: Investigation, sample preparation, writing and editing. MQR: Investigation, review & editing. NS: Investigation, review & editing. NMM: Investigation, writing & review. YMK: Investigation and review. KB: Investigation and review. MR: Investigation and review. JT: Investigation and review. KBJ: Investigation, writing & review. PAD: Investigation and review. ESN: Writing, methodology, data curation, investigation, conceptualization, review & editing, data interpretation, resources, and supervision. GMM: Writing, methodology, investigation, conceptualization, review & editing, data interpretation, and supervision.

## Acknowledgements

We would like to thank the staff members of the Pacific Northwest National Laboratory, a multi-program national laboratory operated by Battelle, Environmental Molecular Sciences Laboratory, for the Department of Energy (DOE) under contract No. DE-AC05-76RLO1830.

## Disclaimer statement

The views and assertions expressed herein are those of the author(s) and do not necessarily reflect the official policy or position of the Department of the Army, Department of Energy, Defense Health Agency, Uniformed Services University, the Department of Defense, or the US Government. The contents of this publication are the sole responsibility of the author(s) and do not necessarily reflect the views, opinions, or policies of The Henry M. Jackson Foundation for the Advancement of Military Medicine, Inc. Mention of trade names, commercial products, or organizations does not imply endorsement by the U.S. Government.

## Supplemental Figures

**Figure S1.**
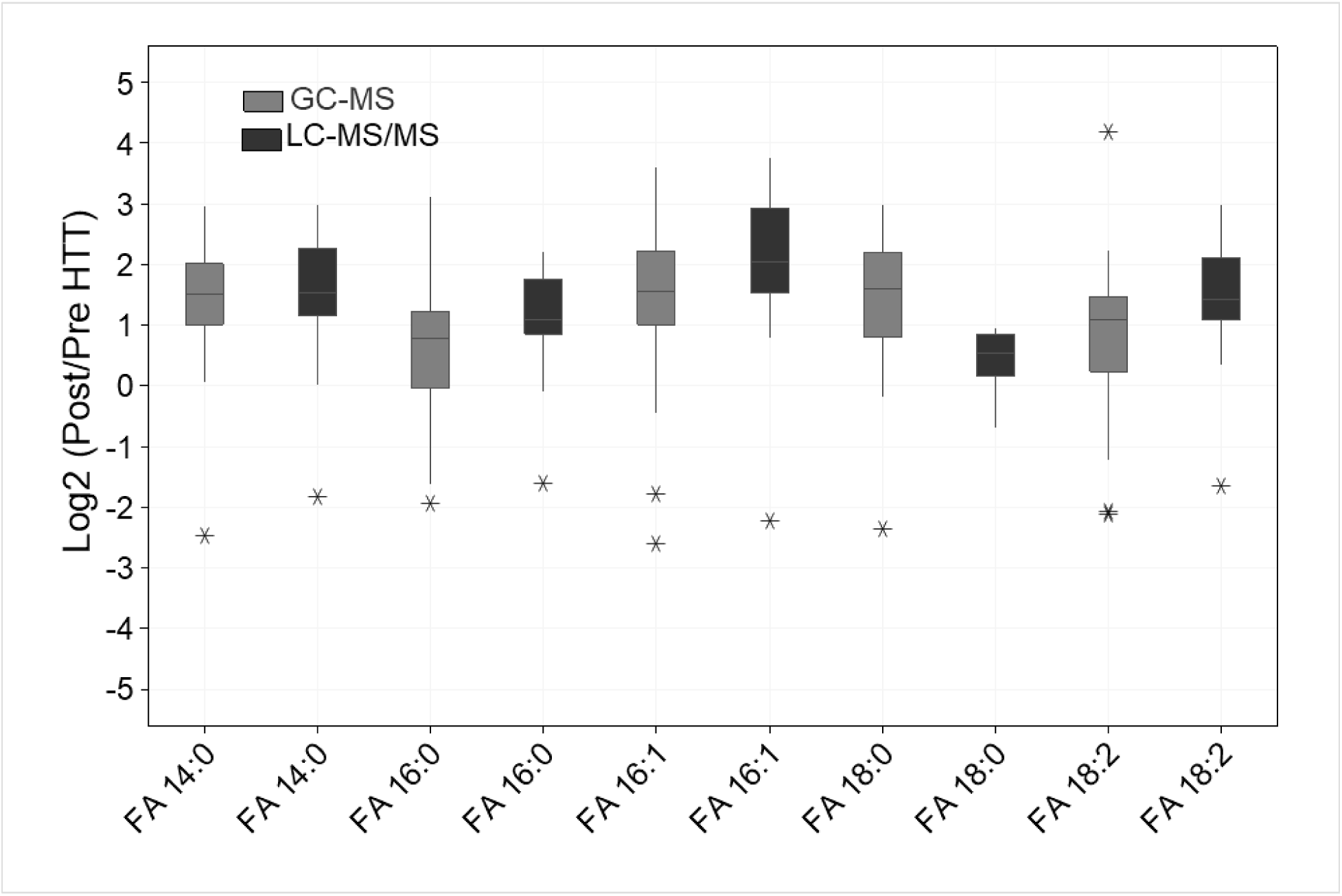
Cross-validation of fatty acids by LC-MS/MS and GC-MS. To confirm the fatty acid profiles observed by liquid chromatography-tandem mass spectrometry (LC-MS/MS), we performed gas chromatography-mass spectrometry (GC-MS) analysis, which showed similar fold change profiles.

**Figure S2.**
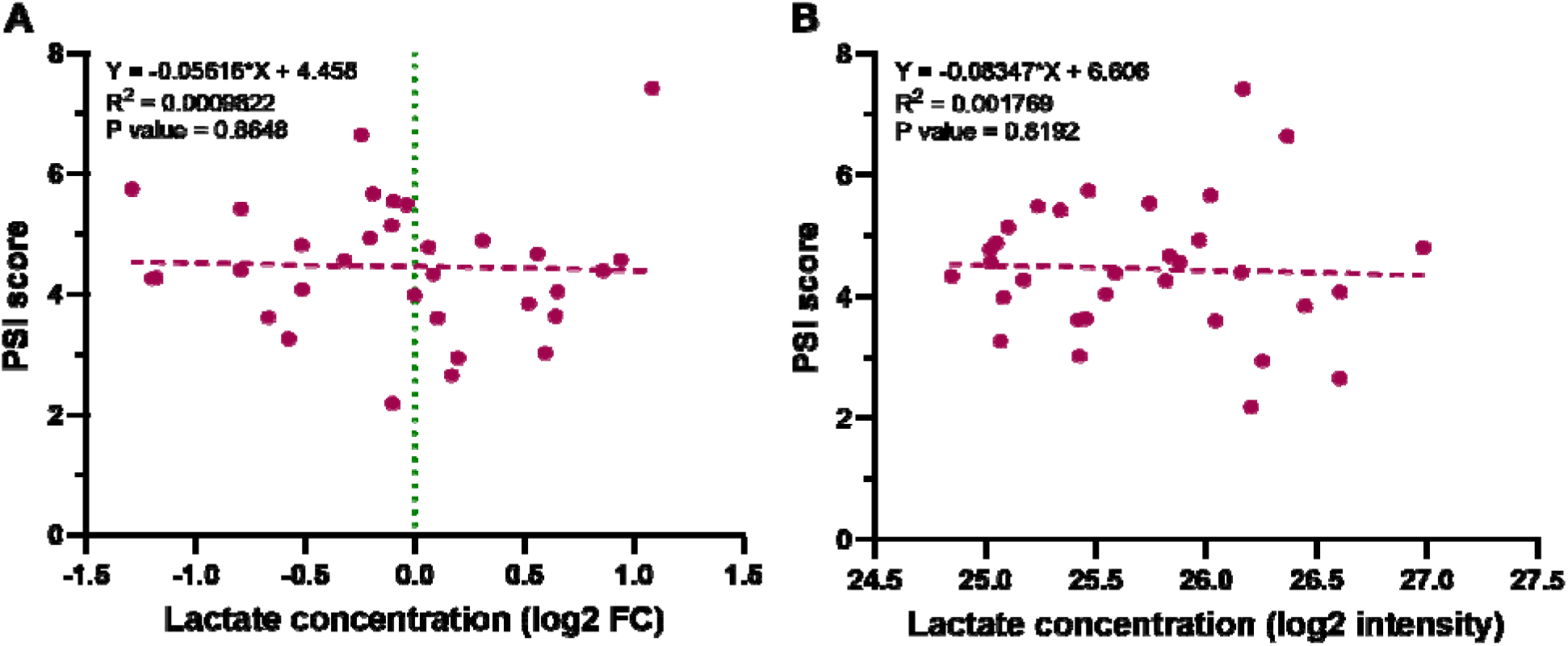
Correlation of heat tolerance score (PSI score) with plasma lactate. **A.** Correlation of PSI score with Post/Pre lactate levels (log2 fold change) in response to the heat tolerance testing. **B.** Correlation of PSI score with the plasma lactate levels immediately following heat tolerance testing.

